# Neutralizing Antibody Responses After mRNA COVID-19 Booster Vaccination are Unaffected by Parasitemia in a Malaria-Endemic Setting

**DOI:** 10.1101/2025.04.12.25325718

**Authors:** Taraz Samandari, Millicent Achola, Jack N. Hutter, Grace Mboya, Walter Otieno, Jia Jin Kee, Yunda Huang, John J. Aponte, Christian F. Ockenhouse, Cynthia K. Lee, Laura Polakowski, Margaret Yacovone, Asa Tapley, Sufia Dadabhai, Nonhlanhla N. Mkhize, Haajira Kaldine, Sinethemba Bhebhe, Penny L. Moore, John Hural, Nigel Garrett, James G. Kublin

**Affiliations:** COVID-19 Prevention Network, Seattle, USA; U.S. Army Medical Research Directorate-Africa, Kisumu, Kenya; Kenya Medical Research Institute, Centre for Global Health Research, Kisumu, Kenya; Vaccine and Infectious Diseases Division, Fred Hutchinson Cancer Center, Seattle, USA; Department of Global Health, University of Washington, Seattle, USA; PATH Center for Vaccine Innovation and Access, Geneva, Switzerland; PATH Center for Vaccine Innovation and Access, Washington, District of Columbia, USA; National Institute of Allergy and Infectious Diseases, National Institutes of Health, Bethesda, USA; Division of Allergy and Infectious Diseases, Department of Medicine, University of Washington, Seattle, USA; Department of Medicine, University of Cape Town, Cape Town, South Africa; Johns Hopkins Bloomberg School of Public Health, Blantyre, Malawi; National Institute for Communicable Diseases of the National Health Laboratory Services, Johannesburg, South Africa; SA MRC Antibody Immunity Research Unit, School of Pathology, Faculty of Health Sciences, University of the Witwatersrand, Johannesburg, South Africa; Desmond Tutu HIV Centre, University of Cape Town, South Africa; Centre for the AIDS Programme of Research in South Africa, University of KwaZulu-Natal, Durban, South Africa; Discipline of Public Health Medicine, School of Nursing and Public Health, University of KwaZulu-Natal, Durban, South Africa

**Author notes:** **Corresponding author** Taraz Samandari, MD PhD, 2520 Kimbolton Dr. College Station, TX 77845.

**Keywords:** mRNA vaccine, malaria, HIV, SARS-CoV-2, booster, immunogenicity, neutralizing antibody

## Abstract

Subclinical malaria may reduce the immunogenicity of mRNA vaccines. We evaluated neutralizing antibody responses in adults with (n=87) and without (n=221) PCR-confirmed *Plasmodium falciparum* who received a COVID-19 booster. Similar boosted ID50 geometric mean titers >22,000 in parasitemic and non-parasitemic participants suggests that COVID-19 mRNA vaccine responses are not impaired.

## INTRODUCTION

mRNA vaccines have demonstrated efficacy against COVID-19 and hold great promise for transforming infectious disease prevention, including for HIV, malaria and tuberculosis.^1^ These diseases are common in the same geographic areas where *Plasmodium falciparum* (Pf) malaria is endemic raising concerns that parasitemia may impair immunity for mRNA vaccines. Diminished vaccine-induced immune responses in parasitemic persons may depend on their age, degree of parasitemia, clinical vs subclinical malaria, and the type of vaccine. Subclinical malaria has been linked to T cell exhaustion and may adversely affect vaccine responses.^2^ To investigate this phenomenon in the context of COVID-19 mRNA vaccines, we conducted a single site substudy of the multisite CoVPN 3008 trial^3^ to assess the immunogenicity of mRNA booster vaccinations in participants with subclinical malaria from Kombewa, Kenya, a region with high seasonal malaria transmission.

## METHODS

The Kombewa site-specific protocol and amendments were approved by Kenya’s Scientific and Ethics Review Unit, with written informed consent obtained from all trial participants before enrollment. The parent trial, CoVPN 3008, was registered on ClinicalTrials.gov, NCT05168813.

The substudy enrolled adults (≥18 years) with HIV (PWH) or other comorbidities associated with severe COVID-19 without exclusions for pregnancy, CD4+ T-cell count, antiretroviral therapy (ART) use, or detectable HIV viral load (VL) who were asymptomatic for COVID-19 or malaria.

Participants had previously received one or two 100-mcg doses of monovalent mRNA-1273, based on whether they were baseline point-of-care SARS-CoV-2 anti-Spike (POC anti-S, Assure Ecotest, Assure Tech, Hangzhou, China) seropositive or seronegative, respectively. During pre-enrolment, participants were randomized 1:1 in a double-blind fashion to receive a booster vaccination with either the monovalent mRNA-1273 or bivalent mRNA-1273.222 (WA-1 and BA.4/5). Further trial details have been published previously.^3^

### Malaria diagnosis

Malaria detection was performed using dried blood spots collected at M0 and M1 and pre-enrolment for most participants (Supplementary Figure 1). Pf was identified via reverse transcriptase polymerase chain reaction (PCR) targeting Pf/pan-Plasmodium 18S rRNA.^4^

### Antibody immunogenicity assay

Serum samples were collected at M0 and M1 and tested at the National Institute for Communicable Diseases, South Africa, using a vesicular stomatitis virus (VSV)-based neutralization assay, which is not affected by ART interference.^5^ Heat-inactivated serum samples underwent five-fold serial dilutions in 96-well plates before addition of the SARS-CoV-2 pseudovirus. The serum-virus complexes were mixed with Vero E6 cells and incubated for 20-24 hours. The SARS-CoV-2 D614G pseudotyped virus was generated by transfecting Expi293F cells with the D614G spike plasmid, along with a VSVΔG plasmid and incubated for 72 hours. Infection was detected via luminescence of the luciferase gene. Neutralizing titers were calculated as the reciprocal serum dilution corresponding to the 50% inhibitory dose (ID50) and 80% (ID80) levels.

In addition to POC anti-S testing, all participants underwent M0 SARS-CoV-2 nasal swab nucleic acid amplification testing (NAAT) and anti-nucleoprotein (anti-NP, Abbott SARS-CoV-2 IgG, Abbott, Chicago, IL, USA) serology testing. Participants with a positive result from any of these tests (nasal swab NAAT, POC anti-S, or anti-NP) were considered to have evidence of prior SARS-CoV-2 infection. Since all participants received at least one dose of COVID-19 mRNA vaccine before enrolment into the malaria substudy, those with prior infection were classified as having “hybrid immunity,” while the remaining participants were classified as having “vaccine immunity.”

### Statistical analyses

The primary objective cohort included participants who (a) received the booster at M0; (b) had malaria PCR test results at both M0 and M1 or were Pf-PCR positive at M0 and missing at M1; (c) had their M1 neutralizing antibody titer collected within 15-42 days post-booster.

Participants with a negative Pf-PCR at M0 and a positive test result at M1 were excluded because their inclusion could alter the response to the booster. In the primary objective cohort analysis, participants were classified as *Pf*-uninfected if Pf-PCR was negative at both M0 and M1, regardless of earlier Pf-PCR results.

We used the SARS-CoV-2 anti-D614G-Spike neutralizing antibody titer as the immunogenicity biomarker, as it is an inverse correlate of risk for symptomatic COVID-19.^6^ The primary endpoints were anti-D614G-Spike-neutralization at the ID50 and ID80 levels at M0 and M1, summarized by geometric mean titers (GMT) and geometric mean fold-rise ratio (GMFR) from M0 to M1. Titers below the lower limit of quantification (LLOQ) were imputed as half the value of LLOQ.

### Malaria substudy sample size

All 330 participants enrolled in CoVPN 3008 at the Kombewa site were invited to join the malaria substudy.

## RESULTS

Participants were enrolled into the substudy and received the booster between January and March 2023, coinciding with seasonal peak malaria activity at the site. All Kombewa participants were PWH. A total of 22 participants were excluded: four due to missing malaria tests at M1, 17 who converted from *Pf*- negative to *Pf* -positive between M0 and M1, and one with missing neutralizing antibody results. The analysis included the remaining 87 *Pf*-positive participants and 221 *Pf*-negative (Supplementary Figure S2).

Among the 308 participants analyzed for the primary objective, 70.8% were female and median age was 37 years (range 20-72). At M0, 7.8% had CD4+ T-cell counts <350 cells/mm^3^, 22.4% had a VL ≥40 copies/mL and 83.8% had evidence of hybrid immunity. Participants with malaria were more likely to have a body mass index ≤25 (88.5% vs 78.3%, p=0.042) and more likely to have a VL ≥40 copies/mL (29.9% vs 19.5%, p=0.0491) than *Pf*-negative participants (Supplementary Table S1).

Figure 1A shows a statistically non-significant lower ID50 anti-D614G Spike neutralizing antibody titer response at M0 in participants with vs without malaria, GMT 2,079 and 2,765 (p=0.084). Similarly, at M1 both groups exhibited a robust booster response, GMT 22,019 and 26,932, respectively (p=0.270).

**Figure 1.**
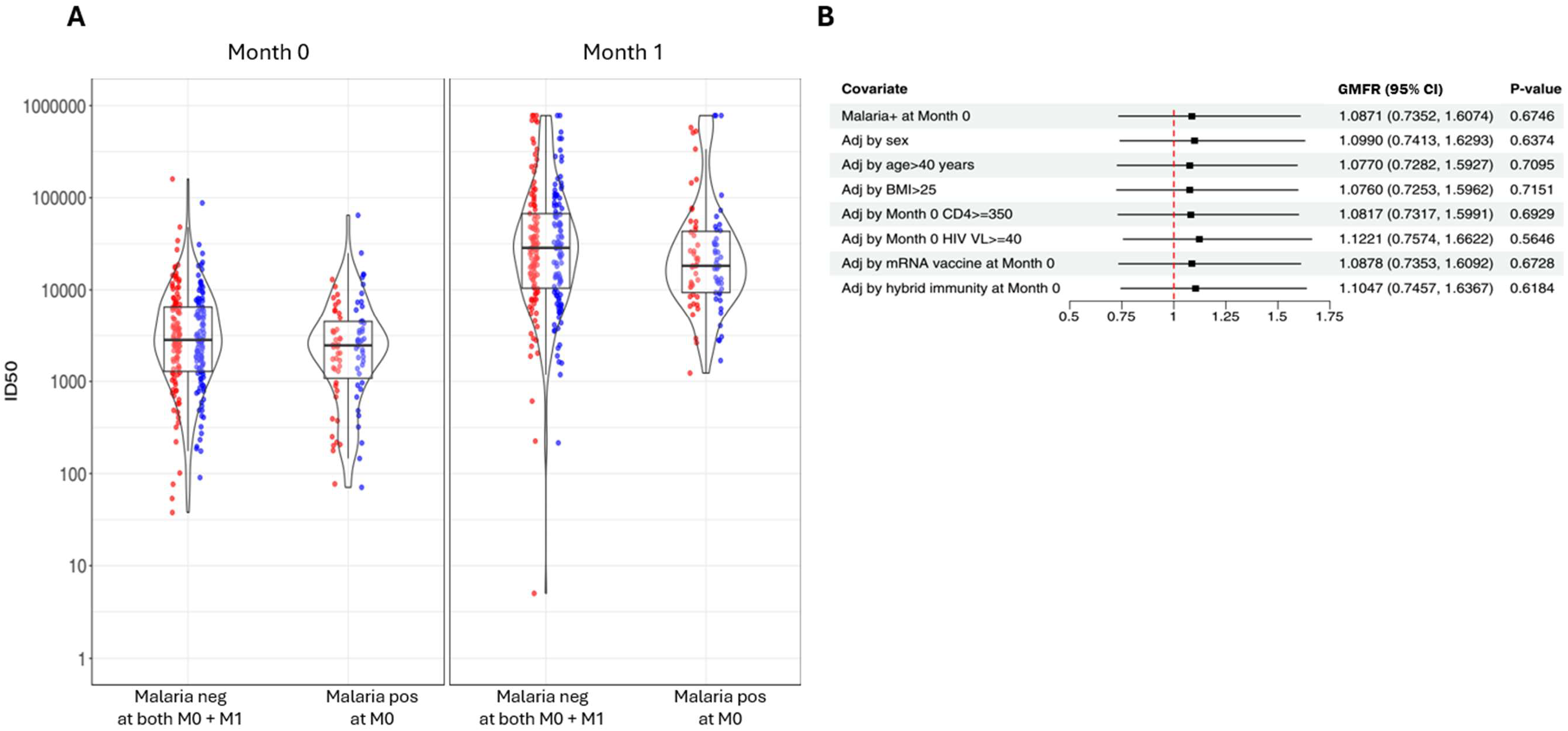
*A*, Violin boxplots of ID50 neutralizing anti-D614G Spike antibody to booster vaccines in asymptomatic Pf-PCR-positive and -negative participants at M0 and M1 (red=monovalent mRNA-1273, blue=bivalent mRNA-1273.222. *B*, ID50 GMFR, M1 over M0, comparing participants with and without malaria and adjusted for covariates in univariate analysis. Abbreviations: Adj, adjusted; BMI, body mass index; CD4, CD4+ T cells measured at M0; CI, confidence intervals; GMFR, geometric mean fold-rise ratio; ID50, 50% inhibitory dilution neutralizing anti-SARS-CoV-2 antibody titer; M0, Month-0 which was the time of booster receipt; M1, Month-1 after booster receipt; neg, Pf-PCR negative; Pf, *Plasmodium falciparum*; pos, Pf-PCR positive; VL, HIV viral load measured at M0.

The ID50 GMFR was comparable between participants with malaria (10.6, 95% confidence interval [CI] 7.6-14.8) and without malaria (9.7, 95% CI 7.9-12.0, Supplementary Table S2). The GMFR between *Pf*- positive and *Pf*-negative was not significantly different, 1.1 (p=0.675), and none of the following covariates significantly affected this relationship: sex, age >40 years, body mass index >25, VL ≥40 copies/mL at M0, monovalent vs bivalent booster or hybrid vs vaccine immunity at M0 (Figure 1B).

Additional analyses were conducted for ID50 and ID80 neutralizing anti-D614G-Spike antibody levels and using alternative definitions of malaria status: (a) considering participants as Pf-positive if they tested positive at either M0 or M1; (b) excluding Pf-negative participants if they had tested positive 4-5 months prior to substudy enrolment; (c) excluding Pf-negative participants if they had tested positive up to 6 months before enrolment. These additional analyses had no impact on the overall findings (see Supplementary Figure 3 and Tables S3-7).

## DISCUSSION

Given the potential importance of mRNA vaccines for preventing infectious diseases in malaria-endemic settings, we sought to determine whether parasitemia adversely affected the immune correlate of protection for COVID-19^6^ following booster vaccination. In this cohort of PWH, we found no reduction in the neutralizing anti-Spike antibody responses to a booster dose of either the monovalent mRNA-1273 or the bivalent mRNA-1273.222 vaccine. Immunogenicity was also unaffected by various potentially confounding factors.

Studies of vaccines in parasitemic children and adults have shown mixed effects on vaccine immunogenicity.^7,8^ These studies included recombinant protein, polysaccharide, and viral vector vaccines. By examining the immunogenicity of a COVID-19 mRNA vaccine in asymptomatic *Pf*-positive adults, our study makes novel contributions to this literature.

Notably, our findings align with recent studies of parasitemic adults immunized with Ebola virus vaccines (rVSVΔG-ZEBOV-GP and Ad26.ZEBOV, MVA-BN-Filo regimen), children immunized with the anti-malaria vaccine RTS,S/AS01 and adults vaccinated with RTS,S/AS01_E_, none of which reported reductions in antibody responses.^9-12^ Immune responses may vary according to specific vaccine platforms and recipient characteristics, emphasizing the need to determine actual vaccine efficacies in *Pf*-positive persons rather than relying entirely on immunogenicity. In the case of COVID-19, the correlate of protection has been established and was utilized in our evaluation.

Our study has some limitations. First, repeated SARS-CoV-2 infections among participants prior to receiving the booster vaccination could have masked reductions in immunogenicity due to parasitemia. Second, a larger sample size may have improved the precision of our GMFR estimates. Third, our use of a 100 mcg booster dose may have induced a higher antibody response than the currently recommended 50 mcg booster dose; leaving uncertainty about the effect of the approved lower dose of the mRNA vaccine in parasitemic individuals. Lastly, parasitemia was measured qualitatively and not quantified preventing a more detailed evaluation of immune responses based on the level of parasitemia. As asymptomatic parasitemic persons typically have low levels of circulating protozoa, this latter issue may not be a significant concern.

In summary, among PWH, the antibody response to mRNA booster vaccination was comparable between those with and without subclinical malaria, supporting the further deployment of mRNA vaccines in malaria-endemic regions.

## Data Availability

This study accesses data through the HIV Vaccine Trials Network (HVTN). Permission to access data will have toshould be requested from HVTN and the Statistical Center for HIV/AIDS Research & Prevention (SCHARP).

## NOTES

## Acknowledgements

The authors thank the participants and study staff of the CoVPN 3008 study.

## Disclaimer

The analyses, conclusions, opinions, and statements expressed herein are solely those of the authors and do not reflect those of the funding or data sources; no endorsement is intended or should be inferred.

## Data Availability

This study accesses data through the HIV Vaccine Trials Network (HVTN). Permission to access data should be requested from HVTN and the Statistical Center for HIV/AIDS Research & Prevention (SCHARP).

