## Supplementary Figures and Tables for "Neutralizing Antibody Responses After mRNA COVID-19 Booster Vaccination are Unaffected by Parasitemia in a Malaria-Endemic Setting"

Supplementary Figure S1. Activities during parent study (“Pre-enrolment”) in relation to the Kombewa malaria substudy (“Enrolment”)

| | Pre-enrolment | | | | | | Enrolment | |
|---|---|---|---|---|---|---|---|---|
| | M0 | M1 | M2 | M3 | M4 | M5 | M0 | M1 |
| Group 1 SARSC2-* (n=121) | 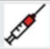 | 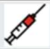 | | | | | 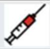 † | |
| Group 2 SARSC2+* (n=205) | 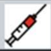 | | | | | | 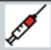 † | |
| *Pf* RT-PCR | 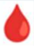 ‡ | 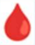 ** | | | | | 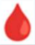 | 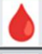 |
| anti-Spike neut antibody | | | | | | | 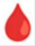 | 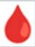 |

NOTE

* 4/330 participants were excluded due to missing malaria tests at Enrolment Month 0

† Randomized 1:1 at Pre-enrolment Month 0 to receive either mRNA-1273 or mRNA-1273.222

‡ Since the parent study (Pre-enrolment period) began enrolment before the malaria sub-study was IRB approved, not all participants had a Pre-enrolment M0 *P falciparum* (Pf-PCR)

** Since Group 1 participants had a baseline negative point-of-care anti-SARS-Cov-2 (SARSC2) antibody test, they returned at Pre-enrolment M1 to receive a second vaccine and were re-tested with Pf-PCR

Supplementary Figure S2. CONSORT diagram showing parent cohort and Kombewa substudy cohort

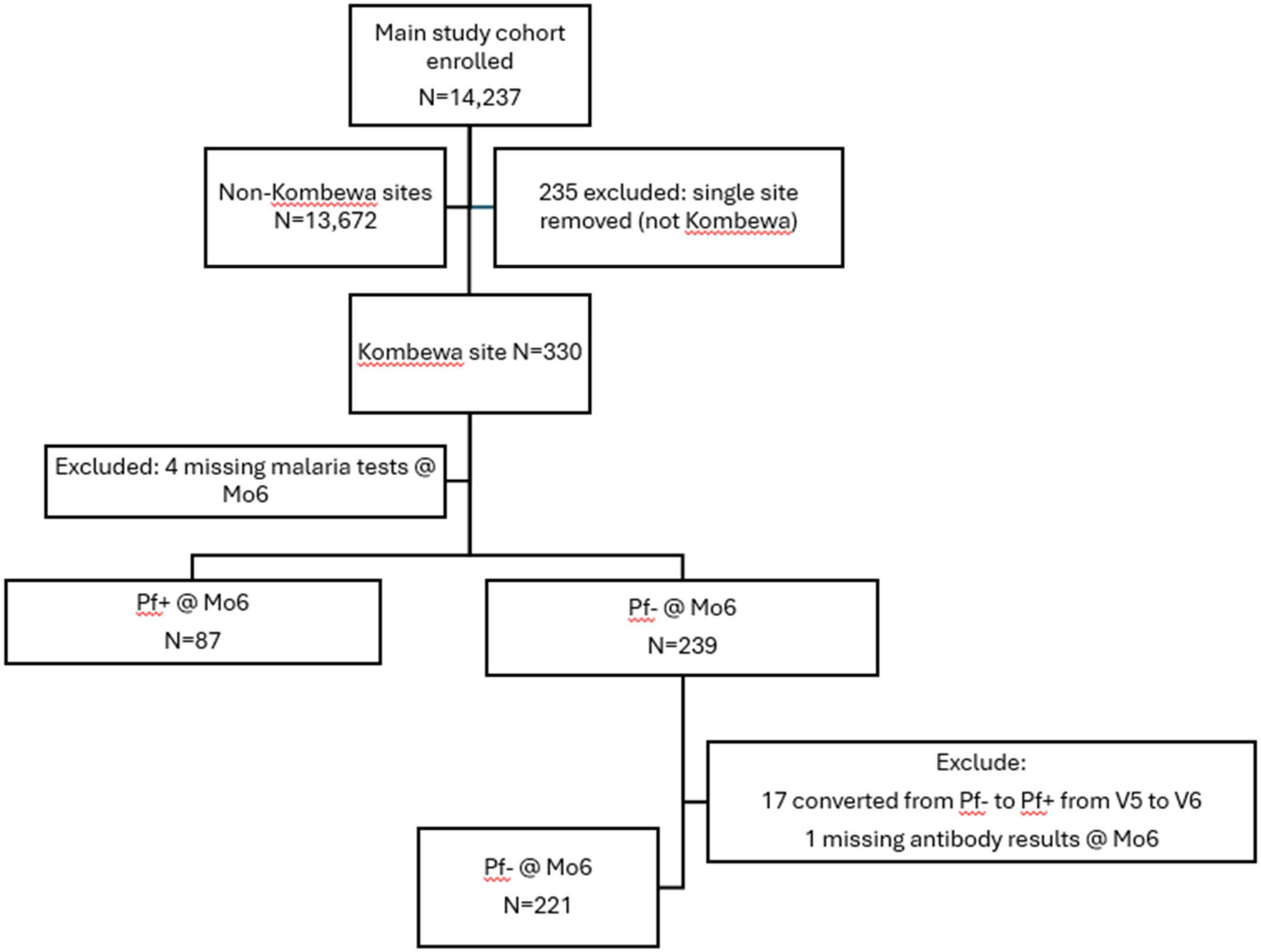

Supplementary Table S1. Baseline characteristics at enrolment (M0) in asymptomatic Pf-PCR-positive (Malaria+) and -negative (Malaria-) participants using the primary objective cohort

| Characteristic | Malaria+ at V5, N = 87 | Malaria- at V5 and V6, N = 221 | Total, N = 308 | P-value* |
|---|---|---|---|---|
| Sex at birth - N (%) | | | | 0.0680 |
| Female | 55 (63.2%) | 163 (73.8%) | 218 (70.8%) | |
| Male | 32 (36.8%) | 58 (26.2%) | 90 (29.2%) | |
| Age category - N (%) | | | | 0.4840 |
| <=40 years | 52 (59.8%) | 142 (64.3%) | 194 (63.0%) | |
| >40 years | 35 (40.2%) | 79 (35.7%) | 114 (37.0%) | |
| BMI at baseline (kg/m2) - N (%) | | | | 0.0421 |
| <=25 | 77 (88.5%) | 173 (78.3%) | 250 (81.2%) | |
| >25 | 10 (11.5%) | 48 (21.7%) | 58 (18.8%) | |
| CD4 at V5 (cells/mm3) - N (%) | | | | 1.0000 |
| <350 | 6 (6.9%) | 18 (8.1%) | 24 (7.8%) | |
| >=350 | 81 (93.1%) | 203 (91.9%) | 284 (92.2%) | |
| HIV viral load at V5 (copies/mL) - N (%) | | | | 0.0491 |
| <40 | 61 (70.1%) | 178 (80.5%) | 239 (77.6%) | |
| >=40 | 26 (29.9%) | 43 (19.5%) | 69 (22.4%) | |
| Month 6 vaccination - N (%) | | | | 1.0000 |
| mRNA 1273 | 43 (49.4%) | 111 (50.2%) | 154 (50.0%) | |
| mRNA 1273.222 | 44 (50.6%) | 110 (49.8%) | 154 (50.0%) | |
| Month 6 immunity status - N (%) | | | | 0.1016 |
| Hybrid | 68 (78.2%) | 190 (86.0%) | 258 (83.8%) | |
| Vaccine | 19 (21.8%) | 31 (14.0%) | 50 (16.2%) | |
| Diabetes at baseline - N (%) | 0 (0.0%) | 0 (0.0%) | 0 (0.0%) | 1.0000 |
| Chronic kidney disease at baseline - N (%) | 0 (0.0%) | 0 (0.0%) | 0 (0.0%) | 1.0000 |
| Cancer at baseline - N (%) | 0 (0.0%) | 0 (0.0%) | 0 (0.0%) | 1.0000 |
| Non-HIV immunocompromised state (weakened immune system) or solid organ transplant at baseline - N (%) | 0 (0.0%) | 1 (0.5%) | 1 (0.3%) | 1.0000 |
| Autoimmune disease at baseline - N (%) | 0 (0.0%) | 0 (0.0%) | 0 (0.0%) | 1.0000 |
| Immunodeficiency at baseline - N (%) | 0 (0.0%) | 0 (0.0%) | 0 (0.0%) | 1.0000 |
| Pregnancy at baseline** - N (%) | | | | 0.3797 |
| Yes | 0 (0.0%) | 3 (1.8%) | 3 (1.4%) | |
| No | 55 (100.0%) | 160 (98.2%) | 215 (98.6%) | |

*Note:*
*P-values for comparison between malaria+ and malaria- participants are obtained from Barnard's test.
**Pregnancy at baseline is calculated among females assigned at birth only.
Primary objective cohort: malaria+ if PCR+ at V5; malaria- if PCR- at V5 and V6; ppts not assigned malaria+ or malaria- are excluded.
Participants meet all the following criteria:
1. Received the booster shot at V5 (Month 6).
2. The neutralizing antibody titer was tested at both V5 and V6 and the V6 draw was +/- 14 days of V6, where V6 is 28 days after V5. In other words, if the V6 titer was drawn <15 days or >42 days after the booster the participant will be excluded.

Supplementary Table S2. ID50 and ID80 geometric mean titers of neutralizing anti-D614G Spike antibody to booster vaccines at M0 (booster receipt) and M1 in asymptomatic Pf-PCR-positive (Malaria+) and -negative (Malaria-) participants using the primary objective cohort

| Isolate | Dilution | Visit | Malaria Positivity | N | Geometric Mean Titer (95% CI) | Median (Q1, Q3) | (Min, Max) | P-value* |
|---|---|---|---|---|---|---|---|---|
| SARS-Cov-2 D614G | 50 | 5 | Malaria- at V5 and V6 | 221 | 2765.3 (2331, 3280.5) | 2821.3 (1283.1, 6501.8) | (37.7, 159969.3) | 0.0844 |
| | | | Malaria+ at V5 | 87 | 2079.7 (1576.8, 2743) | 2462.9 (1088.6, 4481.9) | (70.8, 64569.1) | |
| | | 6 | Malaria- at V5 and V6 | 221 | 26931.8 (21778, 33305.1) | 28586 (10423.3, 67174.1) | (5, 781250) | 0.2704 |
| | | | Malaria+ at V5 | 87 | 22019.1 (16443.6, 29485) | 18256.4 (9346.4, 43141.5) | (1231.4, 781250) | |
| | 80 | 5 | Malaria- at V5 and V6 | 221 | 763.4 (637.5, 914.2) | 826.5 (360.7, 1953.1) | (5, 69596.2) | 0.0549 |
| | | | Malaria+ at V5 | 87 | 545.6 (406.9, 731.7) | 563.8 (278.4, 1375.1) | (19, 22253.6) | |
| | | 6 | Malaria- at V5 and V6 | 221 | 7730.7 (6270.9, 9530.3) | 8146.5 (3120.6, 21474.4) | (5, 781250) | 0.1029 |
| | | | Malaria+ at V5 | 87 | 5823.9 (4440.7, 7637.9) | 5117.3 (2965, 9320.7) | (365.1, 150863.3) | |

*Note:*
*P-values for comparison of geometric means between malaria+ and malaria- participants are obtained from T-tests performed on log-transformed titer values.
95% confidence intervals are calculated using Student's t-distribution.

Supplementary Figure S3. Violin boxplots of ID80 neutralizing anti-D614G Spike antibody to booster vaccines in asymptomatic Pf-PCR-positive and -negative participants at M0 and M1 (red=monovalent mRNA-1273, blue=bivalent mRNA-1273.222.

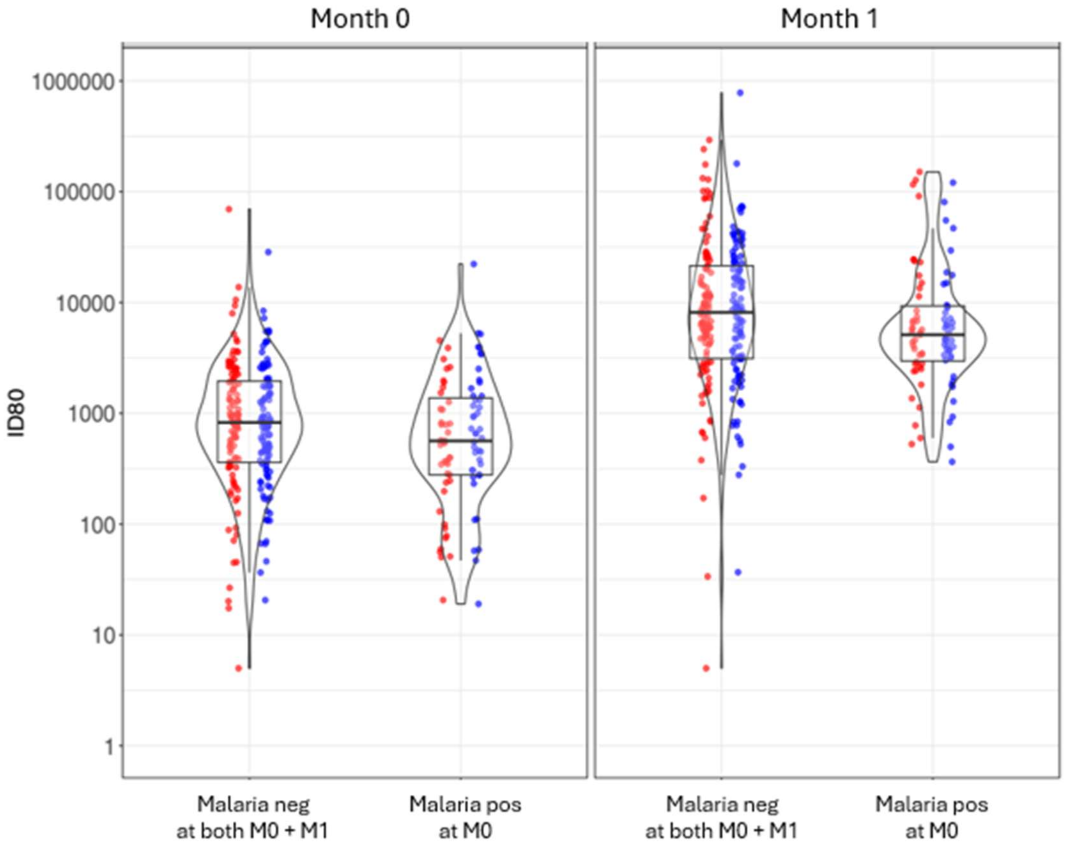

Supplementary Table S3. ID50 and ID80 geometric mean fold rise (M1 over M0) neutralizing anti-D614G Spike antibody to booster vaccines in asymptomatic Pf-PCR-positive (Malaria+) and -negative (Malaria-) participants using the primary objective cohort

| Isolate | Dilution | Malaria Positivity | N | Geometric Mean Fold Rise (95% CI) | Median (Q1, Q3) | (Min, Max) | P-value* |
|---|---|---|---|---|---|---|---|
| SARS-Cov-2 D614G | 50 | Malaria- at V5 and V6 | 221 | 9.7 (7.9, 12) | 8.6 (3.7, 25.7) | (0.1, 652.4) | 0.6756 |
| | | Malaria+ at V5 | 87 | 10.6 (7.6, 14.8) | 8.4 (4, 22.8) | (0.6, 3263.8) | |
| | 80 | Malaria- at V5 and V6 | 221 | 10.1 (8.3, 12.4) | 9.5 (3.9, 24.9) | (0.1, 427.4) | 0.7851 |
| | | Malaria+ at V5 | 87 | 10.7 (7.7, 14.8) | 8.6 (4, 20.2) | (0.6, 1949.4) | |

*Note:*
*P-values for comparison of geometric mean fold rises between malaria+ and malaria- participants are obtained from T-tests performed on log-transformed fold rise values.
95% confidence intervals are calculated using Student's t-distribution.

Supplementary Table S4. Base and univariate models of ID50 and ID80 geometric mean ratio estimate (M1 over M0) comparing Pf-PCR-positive (Malaria+) to -negative participants using the primary objective cohort

| Outcome | Model | Covariate | Geometric Mean Ratio Estimate (95% CI) | P-value |
|---|---|---|---|---|
| ID50 fold rise | Base | malaria_status_pobj1Malaria+ at V5 | 1.0871 (0.7352, 1.6074) | 0.6746 |
| | Univariate 1 | malaria_status_pobj1Malaria+ at V5 | 1.0990 (0.7413, 1.6293) | 0.6374 |
| | | SEXMale | 0.9018 (0.6107, 1.3316) | 0.6021 |
| | Univariate 2 | malaria_status_pobj1Malaria+ at V5 | 1.0770 (0.7282, 1.5927) | 0.7095 |
| | | AGEGR1_f>40 years | 1.2327 (0.8559, 1.7754) | 0.2600 |
| | Univariate 3 | malaria_status_pobj1Malaria+ at V5 | 1.0760 (0.7253, 1.5962) | 0.7151 |
| | | BMIBL_f>25 | 0.9043 (0.5742, 1.4241) | 0.6632 |
| | Univariate 4 | malaria_status_pobj1Malaria+ at V5 | 1.0817 (0.7317, 1.5991) | 0.6929 |
| | | CD4M6_f>=350 | 1.4901 (0.7728, 2.8732) | 0.2328 |
| | Univariate 5 | malaria_status_pobj1Malaria+ at V5 | 1.1221 (0.7574, 1.6622) | 0.5646 |
| | | HIVVLM6_f>=40 | 0.7383 (0.4830, 1.1285) | 0.1604 |
| | Univariate 6 | malaria_status_pobj1Malaria+ at V5 | 1.0878 (0.7353, 1.6092) | 0.6728 |
| | | TRT03P_fmRNA 1273.222 | 0.9266 (0.6513, 1.3184) | 0.6709 |
| | Univariate 7 | malaria_status_pobj1Malaria+ at V5 | 1.1047 (0.7457, 1.6367) | 0.6184 |
| | | SCOV2M6_fVaccine | 0.8138 (0.5036, 1.3150) | 0.3989 |
| ID80 fold rise | Base | malaria_status_pobj1Malaria+ at V5 | 1.0541 (0.7240, 1.5346) | 0.7829 |
| | Univariate 1 | malaria_status_pobj1Malaria+ at V5 | 1.0619 (0.7275, 1.5500) | 0.7550 |
| | | SEXMale | 0.9322 (0.6411, 1.3556) | 0.7125 |
| | Univariate 2 | malaria_status_pobj1Malaria+ at V5 | 1.0407 (0.7153, 1.5143) | 0.8342 |
| | | AGEGR1_f>40 years | 1.3281 (0.9362, 1.8841) | 0.1113 |
| | Univariate 3 | malaria_status_pobj1Malaria+ at V5 | 1.0516 (0.7199, 1.5360) | 0.7941 |
| | | BMIBL_f>25 | 0.9773 (0.6317, 1.5118) | 0.9174 |
| | Univariate 4 | malaria_status_pobj1Malaria+ at V5 | 1.0490 (0.7207, 1.5270) | 0.8021 |
| | | CD4M6_f>=350 | 1.4670 (0.7809, 2.7561) | 0.2327 |
| | Univariate 5 | malaria_status_pobj1Malaria+ at V5 | 1.0847 (0.7436, 1.5823) | 0.6720 |
| | | HIVVLM6_f>=40 | 0.7595 (0.5052, 1.1418) | 0.1853 |
| | Univariate 6 | malaria_status_pobj1Malaria+ at V5 | 1.0549 (0.7243, 1.5365) | 0.7799 |
| | | TRT03P_fmRNA 1273.222 | 0.9051 (0.6451, 1.2699) | 0.5627 |
| | Univariate 7 | malaria_status_pobj1Malaria+ at V5 | 1.0829 (0.7430, 1.5781) | 0.6777 |
| | | SCOV2M6_fVaccine | 0.7080 (0.4470, 1.1213) | 0.1405 |

*Note:*
Covariates besides malaria status included in univariate models have prevalence >=5% in all categories for the cohort.

Supplementary Table S5. Base and univariate models of geometric mean ratio estimate (M1 over M0 comparing Pf-PCR-positive to -negative participants) using the secondary objective ‘a’ cohort (i.e., considering participants as Pf-PCR-positive if they tested positive at either M0 or M1)

| Outcome | Model | Covariate | Geometric Mean Ratio Estimate (95% CI) | P-value |
|---|---|---|---|---|
| ID50 fold rise | Base | malaria_status_sobj1Malaria+ at V5 or V6 | 0.8534 (0.5901, 1.2341) | 0.3984 |
| | Univariate 1 | malaria_status_sobj1Malaria+ at V5 or V6 | 0.8582 (0.5922, 1.2439) | 0.4183 |
| | | SEXMale | 0.9410 (0.6432, 1.3769) | 0.7536 |
| | Univariate 2 | malaria_status_sobj1Malaria+ at V5 or V6 | 0.8470 (0.5857, 1.2250) | 0.3766 |
| | | AGEGR1_f>40 years | 1.2262 (0.8584, 1.7517) | 0.2614 |
| | Univariate 3 | malaria_status_sobj1Malaria+ at V5 or V6 | 0.8409 (0.5797, 1.2199) | 0.3603 |
| | | BMIBL_f>25 | 0.8657 (0.5535, 1.3539) | 0.5262 |
| | Univariate 4 | malaria_status_sobj1Malaria+ at V5 or V6 | 0.8493 (0.5873, 1.2283) | 0.3845 |
| | | CD4M6_f>=350 | 1.4007 (0.7342, 2.6721) | 0.3055 |
| | Univariate 5 | malaria_status_sobj1Malaria+ at V5 or V6 | 0.8749 (0.6036, 1.2681) | 0.4792 |
| | | HIVVLM6_f>=40 | 0.7863 (0.5203, 1.1882) | 0.2528 |
| | Univariate 6 | malaria_status_sobj1Malaria+ at V5 or V6 | 0.8551 (0.5910, 1.2371) | 0.4050 |
| | | TRT03P_fmRNA 1273.222 | 0.9114 (0.6457, 1.2864) | 0.5968 |
| | Univariate 7 | malaria_status_sobj1Malaria+ at V5 or V6 | 0.8597 (0.5936, 1.2452) | 0.4228 |
| | | SCOV2M6_fVaccine | 0.8867 (0.5535, 1.4207) | 0.6162 |
| ID80 fold rise | Base | malaria_status_sobj1Malaria+ at V5 or V6 | 0.8414 (0.5897, 1.2006) | 0.3401 |
| | Univariate 1 | malaria_status_sobj1Malaria+ at V5 or V6 | 0.8439 (0.5902, 1.2068) | 0.3514 |
| | | SEXMale | 0.9688 (0.6713, 1.3982) | 0.8652 |
| | Univariate 2 | malaria_status_sobj1Malaria+ at V5 or V6 | 0.8331 (0.5842, 1.1881) | 0.3123 |
| | | AGEGR1_f>40 years | 1.3101 (0.9296, 1.8463) | 0.1224 |
| | Univariate 3 | malaria_status_sobj1Malaria+ at V5 or V6 | 0.8338 (0.5825, 1.1935) | 0.3195 |
| | | BMIBL_f>25 | 0.9144 (0.5941, 1.4074) | 0.6833 |
| | Univariate 4 | malaria_status_sobj1Malaria+ at V5 or V6 | 0.8378 (0.5871, 1.1956) | 0.3283 |
| | | CD4M6_f>=350 | 1.3595 (0.7294, 2.5336) | 0.3326 |
| | Univariate 5 | malaria_status_sobj1Malaria+ at V5 or V6 | 0.8622 (0.6029, 1.2329) | 0.4153 |
| | | HIVVLM6_f>=40 | 0.7904 (0.5310, 1.1767) | 0.2458 |
| | Univariate 6 | malaria_status_sobj1Malaria+ at V5 or V6 | 0.8431 (0.5906, 1.2036) | 0.3463 |
| | | TRT03P_fmRNA 1273.222 | 0.9124 (0.6546, 1.2718) | 0.5876 |
| | Univariate 7 | malaria_status_sobj1Malaria+ at V5 or V6 | 0.8549 (0.5986, 1.2210) | 0.3876 |
| | | SCOV2M6_fVaccine | 0.7730 (0.4912, 1.2166) | 0.2649 |

*Note:*
Covariates besides malaria status included in univariate models have prevalence >=5% in all categories for the cohort.

Supplementary Table S6. Base and univariate models of geometric mean ratio estimate (M1 over M0 comparing Pf-PCR-positive to - negative participants) using the secondary objective ‘b’ cohort (i.e., excluding Pf-PCR-negative participants if they had tested positive 4-5 months prior to substudy enrolment)

| Outcome | Model | Covariate | Geometric Mean Ratio Estimate (95% CI) | P-value |
|---|---|---|---|---|
| ID50 fold rise | Base | malaria_status_sobj2Malaria+ at V5 | 1.0675 (0.6989, 1.6306) | 0.7617 |
| | Univariate 1 | malaria_status_sobj2Malaria+ at V5 | 1.0872 (0.7092, 1.6667) | 0.7002 |
| | | SEXMale | 0.8640 (0.5714, 1.3064) | 0.4869 |
| | Univariate 2 | malaria_status_sobj2Malaria+ at V5 | 1.0639 (0.6964, 1.6253) | 0.7738 |
| | | AGEGR1_f>40 years | 1.2065 (0.8138, 1.7888) | 0.3486 |
| | Univariate 3 | malaria_status_sobj2Malaria+ at V5 | 1.0627 (0.6940, 1.6272) | 0.7790 |
| | | BMIBL_f>25 | 0.9410 (0.5678, 1.5595) | 0.8127 |
| | Univariate 4 | malaria_status_sobj2Malaria+ at V5 | 1.0554 (0.6911, 1.6117) | 0.8021 |
| | | CD4M6_f>=350 | 1.5796 (0.7990, 3.1228) | 0.1877 |
| | Univariate 5 | malaria_status_sobj2Malaria+ at V5 | 1.1143 (0.7284, 1.7046) | 0.6166 |
| | | HIVVLM6_f>=40 | 0.6748 (0.4254, 1.0702) | 0.0942 |
| | Univariate 6 | malaria_status_sobj2Malaria+ at V5 | 1.0667 (0.6977, 1.6308) | 0.7648 |
| | | TRT03P_fmRNA 1273.222 | 1.0427 (0.7104, 1.5305) | 0.8302 |
| | Univariate 7 | malaria_status_sobj2Malaria+ at V5 | 1.1127 (0.7284, 1.6998) | 0.6201 |
| | | SCOV2M6_fVaccine | 0.6305 (0.3892, 1.0214) | 0.0608 |
| ID80 fold rise | Base | malaria_status_sobj2Malaria+ at V5 | 1.0099 (0.6700, 1.5223) | 0.9622 |
| | Univariate 1 | malaria_status_sobj2Malaria+ at V5 | 1.0232 (0.6764, 1.5480) | 0.9132 |
| | | SEXMale | 0.9009 (0.6035, 1.3449) | 0.6085 |
| | Univariate 2 | malaria_status_sobj2Malaria+ at V5 | 1.0055 (0.6673, 1.5149) | 0.9792 |
| | | AGEGR1_f>40 years | 1.2778 (0.8730, 1.8702) | 0.2062 |
| | Univariate 3 | malaria_status_sobj2Malaria+ at V5 | 1.0085 (0.6674, 1.5238) | 0.9679 |
| | | BMIBL_f>25 | 0.9810 (0.6013, 1.6003) | 0.9385 |
| | Univariate 4 | malaria_status_sobj2Malaria+ at V5 | 0.9987 (0.6628, 1.5049) | 0.9951 |
| | | CD4M6_f>=350 | 1.5651 (0.8088, 3.0285) | 0.1826 |
| | Univariate 5 | malaria_status_sobj2Malaria+ at V5 | 1.0474 (0.6935, 1.5819) | 0.8251 |
| | | HIVVLM6_f>=40 | 0.7161 (0.4578, 1.1201) | 0.1427 |
| | Univariate 6 | malaria_status_sobj2Malaria+ at V5 | 1.0098 (0.6693, 1.5234) | 0.9628 |
| | | TRT03P_fmRNA 1273.222 | 1.0079 (0.6949, 1.4617) | 0.9669 |
| | Univariate 7 | malaria_status_sobj2Malaria+ at V5 | 1.0633 (0.7067, 1.6000) | 0.7675 |
| | | SCOV2M6_fVaccine | 0.5639 (0.3541, 0.8978) | 0.0160* |

*Note:*
Covariates besides malaria status included in univariate models have prevalence >=5% in all categories for the cohort.

Supplementary Table S7. Base and univariate models of geometric mean ratio estimate (M1 over M0 comparing Pf-PCR-positive to -negative participants) using secondary objective ‘c’ cohort (i.e., excluding Pf-PCR-negative participants if they had tested positive up to 6 months before enrolment)

| Outcome | Model | Covariate | Geometric Mean Ratio Estimate (95% CI) | P-value |
|---|---|---|---|---|
| ID50 fold rise | Base | malaria_status_sobj3Malaria+ at V5 | 1.0501 (0.6724, 1.6399) | 0.8291 |
| | Univariate 1 | malaria_status_sobj3Malaria+ at V5 | 1.0656 (0.6786, 1.6732) | 0.7817 |
| | | SEXMale | 0.9030 (0.5775, 1.4118) | 0.6530 |
| | Univariate 2 | malaria_status_sobj3Malaria+ at V5 | 1.0520 (0.6736, 1.6429) | 0.8229 |
| | | AGEGR1_f>40 years | 1.2348 (0.8160, 1.8684) | 0.3166 |
| | Univariate 3 | malaria_status_sobj3Malaria+ at V5 | 1.0264 (0.6535, 1.6120) | 0.9095 |
| | | BMIBL_f>25 | 0.8386 (0.4991, 1.4090) | 0.5044 |
| | Univariate 4 | malaria_status_sobj3Malaria+ at V5 | 1.0460 (0.6697, 1.6338) | 0.8425 |
| | | CD4M6_f>=350 | 1.4122 (0.7004, 2.8473) | 0.3329 |
| | Univariate 5 | malaria_status_sobj3Malaria+ at V5 | 1.1034 (0.7034, 1.7308) | 0.6671 |
| | | HIVVLM6_f>=40 | 0.6984 (0.4202, 1.1609) | 0.1652 |
| | Univariate 6 | malaria_status_sobj3Malaria+ at V5 | 1.0500 (0.6716, 1.6416) | 0.8297 |
| | | TRT03P_fmRNA 1273.222 | 1.0195 (0.6799, 1.5287) | 0.9251 |
| | Univariate 7 | malaria_status_sobj3Malaria+ at V5 | 1.0511 (0.6689, 1.6516) | 0.8281 |
| | | SCOV2M6_fVaccine | 0.9922 (0.5704, 1.7261) | 0.9778 |
| ID80 fold rise | Base | malaria_status_sobj3Malaria+ at V5 | 1.0066 (0.6478, 1.5639) | 0.9767 |
| | Univariate 1 | malaria_status_sobj3Malaria+ at V5 | 1.0200 (0.6530, 1.5934) | 0.9303 |
| | | SEXMale | 0.9115 (0.5860, 1.4179) | 0.6797 |
| | Univariate 2 | malaria_status_sobj3Malaria+ at V5 | 1.0086 (0.6494, 1.5666) | 0.9693 |
| | | AGEGR1_f>40 years | 1.2706 (0.8440, 1.9128) | 0.2498 |
| | Univariate 3 | malaria_status_sobj3Malaria+ at V5 | 0.9915 (0.6344, 1.5496) | 0.9700 |
| | | BMIBL_f>25 | 0.8903 (0.5329, 1.4875) | 0.6558 |
| | Univariate 4 | malaria_status_sobj3Malaria+ at V5 | 1.0025 (0.6452, 1.5576) | 0.9912 |
| | | CD4M6_f>=350 | 1.4381 (0.7192, 2.8757) | 0.3025 |
| | Univariate 5 | malaria_status_sobj3Malaria+ at V5 | 1.0428 (0.6675, 1.6291) | 0.8531 |
| | | HIVVLM6_f>=40 | 0.7735 (0.4675, 1.2797) | 0.3157 |
| | Univariate 6 | malaria_status_sobj3Malaria+ at V5 | 1.0066 (0.6472, 1.5657) | 0.9765 |
| | | TRT03P_fmRNA 1273.222 | 0.9814 (0.6576, 1.4647) | 0.9264 |
| | Univariate 7 | malaria_status_sobj3Malaria+ at V5 | 1.0302 (0.6594, 1.6095) | 0.8956 |
| | | SCOV2M6_fVaccine | 0.8265 (0.4784, 1.4277) | 0.4926 |

*Note:*
Covariates besides malaria status included in univariate models have prevalence >=5% in all categories for the cohort.
